# Quantifying Respiratory Airborne Particle Dispersion Control Through Improvised Reusable Masks: The Physics of Non-Pharmaceuptical Interventions for Reducing SARS-COV-2 (COVID-19) Airborne Transmision

**DOI:** 10.1101/2020.07.12.20152157

**Authors:** Nathan J. Edwards, Rebecca Widrick, Richard Potember, Mike Gerschefske

## Abstract

**Background:** For much of the SARS-CoV-2 (COVID-19) pandemic, many countries have struggled to offer definitive guidance on the wearing of masks or face coverings to reduce the highly infectious disease transmission resulting from a lack of compelling evidence on the effectiveness of communities wearing masks, and slow acceptance that aerosols are a primary SARS-CoV-2 disease transmission mechanism. Recent studies have shown that masks have been effective in several countries and populations, leaving only a lack of quantitative data on the control of airborne dispersion from human exhalation. This current study specifically has the objective to quantify the effectiveness of non-medical grade washable masks or face coverings in controlling airborne dispersion from exhalation (both droplet and aerosol) by measuring changes in direction, particle cloud velocities, and concentration.

**Design:** This randomized effectiveness study used a 10% NaCl nebulized polydisperse particle solution (0.3 μm up to 10 μm in size) delivered by an exhalation simulator to conduct 94 experiment runs with combinations of 8 different fabrics, 5 mask designs, and airflows for both talking and coughing. Multiple particle sensors were instrumented to measure reduction in aerosol dispersion.

**Results:** Three-way multivariate analysis of variance establishes that fabric, mask design, and exhalation breath level have a statistically significant effect on changing direction, reducing velocity, or concentrations of airborne particles (Fabric: *P* = < .001, Wilks’ Λ = .000; Mask design: *P* = < .001, Wilks' Λ = .000; Breath level: P = < .001, Wilks' Λ = .004). There were also statistically significant interaction effects between combinations of all primary factors.

**Conclusions and Relevance:** The application of facial coverings or masks can significantly reduce the airborne dispersion of aerosolized particles from exhalation by diffusing the particle cloud direction and slow down its travel speed. Consequently, the results indicate that wearing masks when coupled with social distance can decrease the potentially inhaled dose of SARS-CoV-2 aerosols or droplets especially where infectious contaminants may exist in shared air spaces. The conclusion is well aligned with the concept of “time-distance-shielding” from hazardous materials emergency response. However, the effectiveness varies greatly between the specific fabrics and mask designs used.

## BACKGROUND

In light of the current pandemic from rapid transmission of the severe acute respiratory syndrome coronavirus 2 (SARS-CoV-2 or COVID-19) and significant morbidity, there has been inconsistent medical guidance given to the public regarding the wearing of non-medical improvised fabric masks or face coverings to reduce the transmission of COVID-19. If the SARS-CoV-2 aerosol is considered with an ability to infect for more than 3 hours with TCID_50_ of greater than 10^2^ as noted in a laboratory study[1] then it is of significant importance for broad public health infectious disease strategy to understand the effectiveness of non-medical masks and face coverings to control human exhalation aerosol dispersion, especially with asymptomatic or pre-symptomatic populations.

Of concern, recent studies show that bio-droplets of all sizes are generated from normal exhalation[2-5] with 80-90% of droplets from human exhalation in the size range of 0.1-1μm,[6, 7] those from a cough can travel 23 to 27 feet (7-8 m) which is well beyond the recommended social distances of six feet or two meters, and smaller aerosols (≤5μm) stay aloft in the air and pose a greater risk for severe infection.[8, 9] In addition, social distancing is difficult to accomplish since many essential locations like grocery stores have aisles that are narrow and result in the proximity of patrons being closer than 1 meter. Reduced distance correlates to increased transmission of COVID-19.[10, 11]

A lack of definitive data on establishing the effectiveness of using non-medical masks or face coverings has resulted in medical practitioners giving broad public health guidance based on professional judgement only. Existing guidance includes statements that facial coverings may offer minimal protection from small infectious particles, may only reduce large particulate matter, or only remind users to not touch their face considering infectious disease transmission from hand to face. One key challenge has been the slow acceptance of aerosol transmission as the primary SARS-CoV-2 disease transmission mechanism.[9, 12] A number of previous studies have been conducted to understand if wearing of masks reduce community infections of common diseases such as influenza, however most are inconclusive due to the application of masks post-exposure or lack of strict wearing compliance by study participants.[13-15] Only a few well-executed studies concluded the prophylactic wearing of medical grade masks reduce community transmission of influenza or RSV.[16, 17] To make matters worse, the lack of definitive guidance has also led to social and political debates on the wearing of masks or face coverings[18-20] and deters the acceptance of any new public health strategy for reducing airborne transmission of infectious diseases.

Only recently, several definitive case studies and broad systematic or meta-analysis shown that masks and face coverings have been effective in reducing community transmission of COVID-19 within several countries and populations,[21-25] which leaves only a lack of quantitative data on the control of airborne dispersion from human exhalation. Prior studies have established the filtration efficiency of a variety of fabrics for personal protection but do not consider reducing transmission by controlling airborne dispersion of human exhalation.[26-33] Other research investigates only forward dispersion of particles from coughing or sneezing by measuring from a single optical plane.[8, 34] Other research considers the aerodynamics of exhalation particles inside various rooms or vehicles using high fidelity computational fluid dynamic simulation under ideal laminar flow conditions and possible effectiveness of masks, [35-39] but do not model real world turbulent airflows nor validate the models with physical experimentation data. Several clinical studies show reductions in virus shedding when wearing face masks,[2, 3, 5, 40-42] although neither the clinical nor experimental studies have fully characterized the effectiveness of non-medical grade reusable masks in controlling aerosol dispersion of human exhalation particles.

## OBJECTIVE, DESIGN, AND METHODS

The goal of this research is to determine the statistically significant factors and quantify the effectiveness of non-medical grade washable masks or face coverings in the control of aerosol dispersion from human exhalation in terms of affecting direction, velocity, and concentration of various particle diameters. When considering the primary contribution of this work, there are three complimentary types of studies regarding the wearing of masks and face coverings in the context of respiratory transmission of infectious disease: mask filtration studies, clinical studies, and particle dispersion studies. Existing mask filtration studies help answer the question of will they protect from inhalation. Clinical studies on masks help answer the question of do they work for broad community reductions in transmission. Particle dispersion studies offer significant insights to the aerodynamics and bio-physics of infectious particles, although there is not as much research in this category. All three types of studies are necessary to inform and construct the most holistic solutions for public health.[43] This original research offers the experimental results in the latter category of aerodynamics and bio-physics of infectious airborne particles. Conclusively this study can aid in establishing public health strategies or policies that encourage the wearing of masks or face coverings to increase the effectiveness of non-pharmaceutical interventions (NPI) especially where infectious contaminants may exist in shared air spaces.

### Design

We conducted the effectiveness study using a randomized full factorial design of experiments with 10% NaCl nebulized solution and an exhalation simulator to conduct 94 experiment runs with combinations of 8 different fabrics, 5 mask designs, no-mask as a control, and exhalation airflows (PEF and FEV1) that represent both talking and coughing. Randomization was created using a mathematical function in MATLAB with blocking on the two airflow settings (47 each). The experiment also included randomized runs of no-mask applied as the control and a preliminary comparison with the performance of a MERV13 air filter media which has similar electrostatic filtration properties to the NIOSH N95 standard (95% filtration efficiency of 0.3μm particles).

The exhalation simulator was constructed similar to previous research,[44, 45] but with some differences. The exhalation simulator was driven by a dry compressed air expansion chamber and timing-controlled relay, with a port for the small volume jet nebulizer, in-line spirometer, and a corrugated tube to emulate a trachea before exiting the mouth of the CPR manikin. Details on the simulator equipment are in (online Supplementary Figure 1). The exhalation airflows were calibrated to simulate peak expiratory flow (PEF) of coughing with a range 507L/min to 650L/min, and PEF for talking of approximately 120L/min as established by previous research.[44-47] The typical air flows for talking are similar to that of singing.[47]

Four laser scattering particle concentration sensors (Plantower PMS5003) that measured particles from 0.3μm to 10μm in size were placed at specific locations inside a non-airtight fume hood (online Supplementary Figure 2) to detect aerosol dispersion directly downward, laterally from the mid-line, and 1 meter forward of the mouth. Preliminary testing of the configuration identified that the optimal position when used with various masks in this fume hood was 43 cm below the level of the mouth for all sensors. The frontal sensor represents an approximate halfway point of a 2 meter or 6 feet social distance.

### Interventions and Exposures

A NaCl aqueous solution was selected as a polydisperse test aerosol which is also used as the exposure for NIOSH N95 respirator test methods. 10% NaCl was used to generate a sufficient quantity of particles for the open-air fume hood environment and also to stay beneath the PMS sensor maximum (65,535 particle count per 0.1L for any given size). The aerosol was produced by nebulizing the solution at 103 kPa (15 psi) for 5 seconds into the aerosol chamber of the exhalation simulator followed by a 3 ms delay before exhalation from the manikin through the applied masks. The simulated exhalations were driven by timing controlled compressed air at 827 kPa (120 psi) for “coughing” and 206 kPa (30 psi) for “talking”.

The intervention was provided by non-medical grade washable masks sourced from local materials that were available during the COVID-19 supply chain interruptions. The fabrics were selected are shown in (online Supplementary Table 1) (fabric bolts were unavailable) which included natural fibers, polyesters, and other materials. The mask designs were selected from variety of community-based designs which included a bandana style, surgical mask style, folded no-sew, a simple mask with earloops, and a stylistic mask that had more coverage of the nose. Microscopic images of the fabric weave and fibers were taken (Keyence VHX-S660E) to further understand and explain the results. More details regarding the fabrics and masks designs and the basic test procedure are also included in the (online Supplementary Appendix).

### Outcomes

The primary outcome was to measure any significant reduction in aerosol dispersion velocity, quantity of particles, and change in dispersion direction. Measurements used in this study included peak expiratory flow (PEF), forced expiratory volume (FEV1), as well as aerosol arrival time, time to peak concentration, aerosol velocity, area under curve (AUC) for first minute and last minute as shown in Figure 1. A change in direction from sensor 5, reduction in velocity, or AUC are considered a positive effect. Two novel metrics of Filtration Efficiency Indicator (FEI) and Expiratory Flow Dispersion Factor (EDF) are established in this study to present quantitative values that give relative indicators to the dispersion control performance of non-medical masks using simple and repeatable measurement techniques of the research. A description of all measurements and outcomes are presented in Table 1.

**Figure 1:**
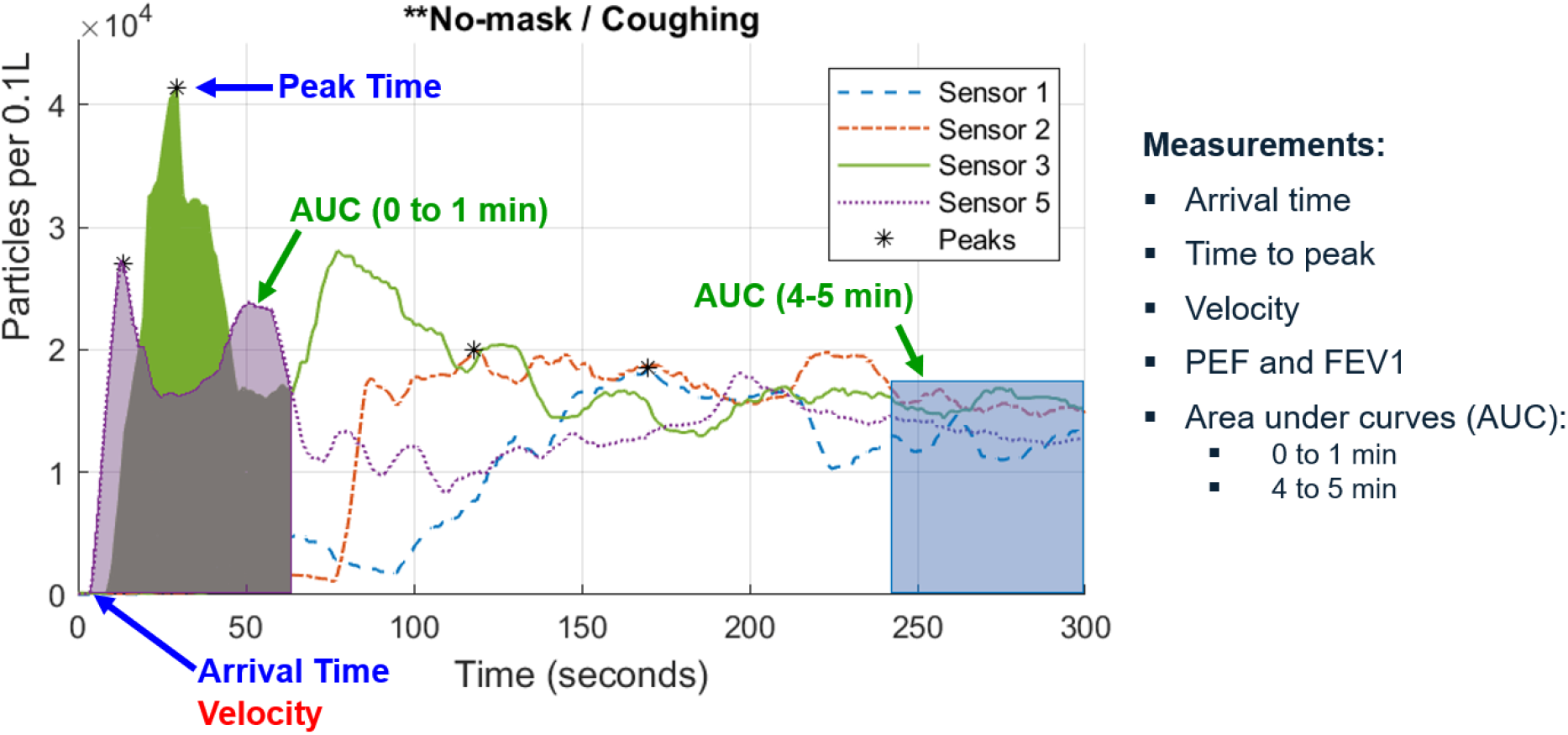
Examples of the measurements performed on the time-series data from all experiment runs for each of the four sensors and particle sizes of 0.3μm, 0.5μm. 1μm, 2.5μm, 5μm, 10μm

**Table 1.**
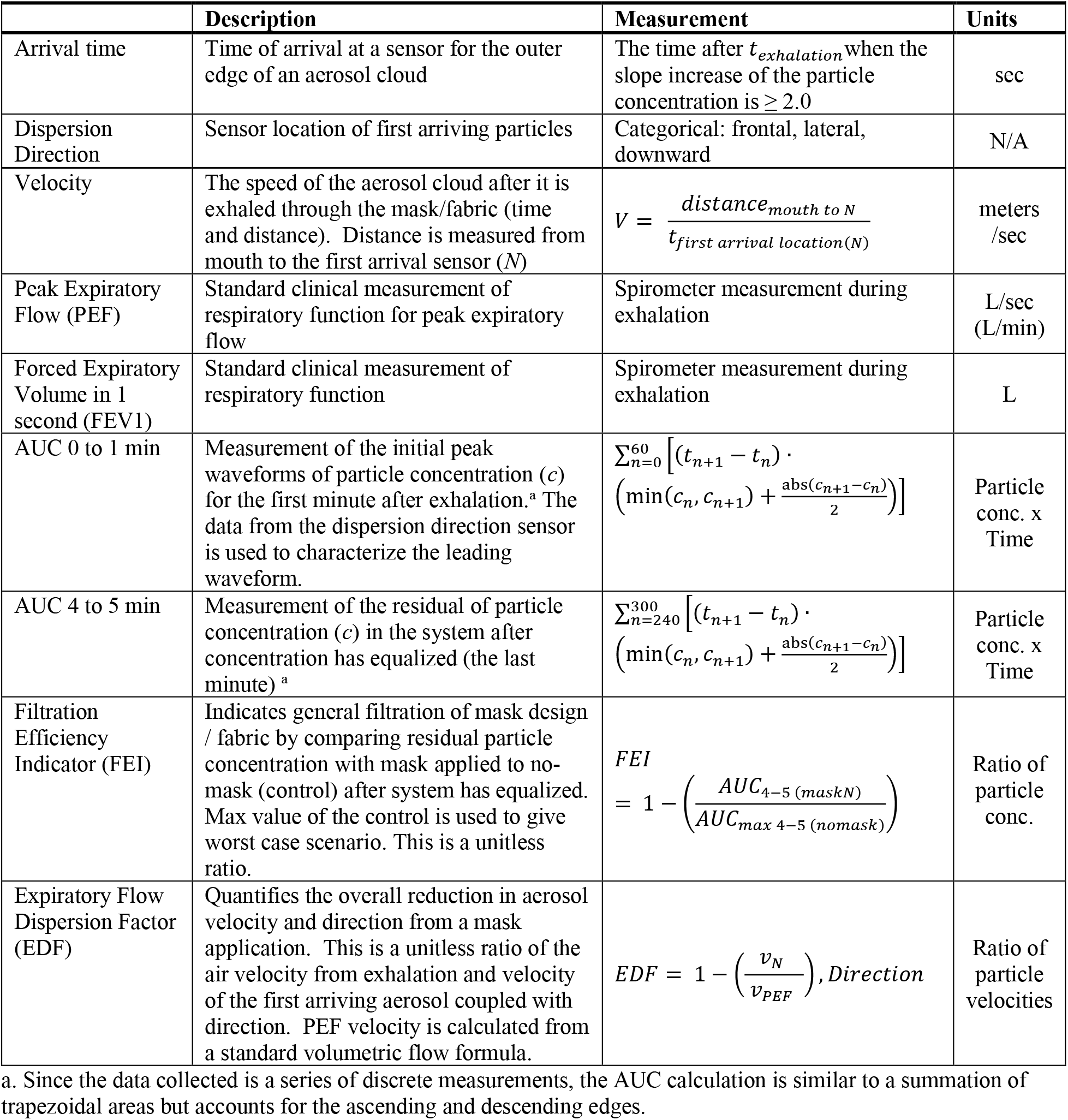
Measured Outcomes and Descriptions

AUC_0 to 1 min_ as described in Table 1 characterizes the leading wave of the particle cloud. A higher value indicates a more particle concentration in the airspace over time. Lower values result from abrupt changes to particle concentration due to an overall lower amount of particles from exhalation or a rapid movement of the particle cloud movement. In general, a lower value is more desirable which represents less time and concentration that a person might be exposed to a particle cloud before it disperses.

FEI is a ratio of the particle concentration remaining after exhalation through a mask compared to no-mask and provides a quantitative indicator that aids in the filtration performance characterization. It is a ratio of remaining particle concentration (AUC_4 to 5 min_) with a mask applied compared to the worst-case AUC of no-mask applied; a higher value indicates better filtration. Since this experiment does not lend itself to directly measure the particle concentration in the aerosol chamber prior to exhalation nor does it use the same measurement equipment, test orifice and tube size, airflow dynamics, and other equipment from the NIOSH N95 test standard,[48] the filtration efficiency indicator values are relative to this experiment.

Likewise, many other recent studies seeking to establish the filtration efficiencies of non-medical grade masks or fabrics have the same constraint on relativity of results. However, while the actual values are relative to this study the FEI measurement technique is broadly applicable to all exhalation dispersion studies.

We also present EDF as a measurement of the reduction in particle velocity and change in direction when a mask is applied. As shown in Table 1, it is the ratio of particle cloud velocity at the first arriving sensor and that of the exhalation airflow velocity derived from the PEF measurement. A higher value indicates better performance with more reduction in particle velocities. The theory of EDF is based on the airflow from exhalation simulator (bounded volume) and aerosol velocity in fume hood (unbounded turbulent airflow) which are correlated by Bernoulli’s ideal-gas law and further described in the field of kinetic theory of gases. Full derivation of the volumetric flow formula is provided in literature;[49] in this study the airflow of PEF equals the cross-sectional area of the spirometer multiplied by the average velocity of the air stream shown in Equation (1)

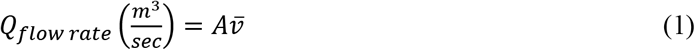

Where:

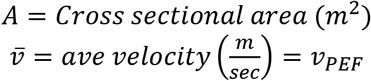

The inside diameter of the MIR SmartOne spirometer was measured to be 28.67 mm which allows for an area calculation. Using algebraic relationships, the formula for calculating the velocity of the exhalation using PEF measurement is shown in Equation (2) along with the unit conversion to meters per second.

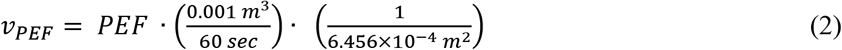

**Sample Size:** The overall sample size from the full factorial combination of 40 distinct masks, several randomly inserted test runs of no-mask as the control, and 2 exhalation levels resulted in 94 experiment runs. The overall experiment with four sensors and average sampling rate of 1 second, generated over 1.694 million time-series sensor data measurements for this study. The sample size of *n*=94 resulted in 2,496 measurements for each dependent response variable across all particle diameters.

### Statistical Analysis

Multivariable and multivariate analysis was conducted from the perspective of a null hypothesis that non-medical improvised masks do not affect the dispersion or offer any source control. This study determines if the three independent variables (mask designs, fabrics and breathing levels) have a statistically significant effect on any of the dependent responses (direction, AUC_0 to 1 min_, FEI, and EDF) at various sensor locations. Three-way multivariate analysis of variance (MANOVA) is used to simultaneously understand the significance of multiple effects from the independent variables and their correlations while minimizing type I statistical errors (false positives). Details on the use of MANOVA and validation against the assumptions of the data, including homogeneity of covariance, normality, independence of observations and multicollinearity[50-54] are provided in (online Supplementary Description of Statistical Methods).

MATLAB version R2019a was used to import the raw data files, compute the response variable values, and calculate summary statistics. SPSS version 1.0.0.1327 was used to perform MANOVA. To additionally validate the statistical results and measured outcomes, graphical analysis of the data was also performed to identify any anomalies that were not expected in the response variables.

## RESULTS

The mean (SD) PEF for simulated coughing was 532.08 (75.65) L/min and FEV1 of 5.92 (0.1) L. Likewise, the mean (SD) PEF for simulated talking was 148.35 (43.29) L/min and FEV1 of 1.79 (0.07) L. Both simulated exhalation levels are within range of previous studies.[44-47] The aerosol particle concentration was measured at the one-meter frontal sensor during the last minute of all no-mask (control) runs and resulted in concentration levels and distribution that indicates good polydisperse particle generation (online Supplementary Figure 3). The mean (SD) concentrations for simulated talking generated peak concentrations of 32,199 (3,683) for 0.3μm particle diameters representing aerosols, and 201(42) for 10μm particle diameters representing droplets. Likewise, simulated coughing generated peak concentrations of 27,731 (9,837) for 0.3μm particle diameters, and 131(27) for 10μm particle diameters.

Table 2 shows descriptive summary statistics of variables with respect to the primary dispersion direction and gives some insight into the generalized responses. The best overall performing mask is the surgical style with internal non-woven layers [AUC_0 to 1 min_ = 7.721×10^5^ (6.606×10^5^), FEI = .468(.158), EDF = .993(.005)]. The best overall fabric depends on a desired characteristic of reduced velocity and direction or increased filtration performance; a general comparison of EDF across mask designs is shown in Figure 2. The velocity-ratio related performance, EDF, indicates the overall slowdown of particles due to turbulence and aerodynamics in an open air system. The large standard deviations represent the divergence between the responses for each exhalation breath level (visible in online Supplementary Figures 6 – 13) and also indicate that the interactions between multiple factors and the multivariate responses.

**Table 2.** Descriptive statistics of variables used in the MANOVA with respect to primary dispersion direction

| Fabric | n | Direction |  | Velocity of Dispersion |  | AUC 0to1 |  | FEI |  | EDF |  |
| --- | --- | --- | --- | --- | --- | --- | --- | --- | --- | --- | --- |
|  |  | Mode | % | Mean | SD | Mean | SD | Mean | SD | Mean | SD |
| **No-mask | 14 | 5 | 100.0% | --<sup>a</sup> | -- | 1.059<br>$\times 10^6$ | 2.237<br>$\times 10^5$ | 0.092 | 0.084 | --<sup>a</sup> | -- |
| Bandana, 100% cotton bandana | 10 | 1 | 100.0% | 0.216 | 0.217 | 1.846<br>$\times 10^6$ | 9.194<br>$\times 10^5$ | 0.409 | 0.167 | 0.962 | 0.045 |
| Bath towel 100% combed cotton | 10 | 3 | 83.3% | 0.170 | 0.213 | 1.314<br>$\times 10^6$ | 7.995<br>$\times 10^5$ | 0.471 | 0.166 | 0.983 | 0.013 |
| Pillowcase, 60% cotton, 40% polyester | 10 | 5 | 66.7% | 0.059 | 0.058 | 1.516<br>$\times 10^6$ | 8.360<br>$\times 10^5$ | 0.352 | 0.200 | 0.992 | 0.005 |
| Sweat pant, 60% cotton, 40% polyester | 10 | 2 | 83.3% | 0.155 | 0.178 | 1.591<br>$\times 10^6$ | 8.081<br>$\times 10^5$ | 0.469 | 0.173 | 0.984 | 0.012 |
| T-shirt, 60% cotton, 40% polyester | 10 | 1 | 100.0% | 0.122 | 0.087 | 1.839<br>$\times 10^6$ | 8.884<br>$\times 10^5$ | 0.444 | 0.109 | 0.982 | 0.012 |
| Dress shirt, 100% polyester | 10 | 5 | 100.0% | 0.100 | 0.166 | 1.298<br>$\times 10^6$ | 9.573<br>$\times 10^5$ | 0.304 | 0.151 | 0.977 | 0.049 |
| Mover blanket, 85% polyester, 15% recycled cotton | 10 | 5 | 100.0% | 0.138 | 0.228 | 1.601<br>$\times 10^6$ | 1.060<br>$\times 10^6$ | 0.331 | 0.099 | 0.966 | 0.066 |
| Microfiber towels, 80% polyester, 20% polyamide | 10 | 1 | 100.0% | 0.090 | 0.079 | 1.382<br>$\times 10^6$ | 8.543<br>$\times 10^5$ | 0.405 | 0.165 | 0.986 | 0.011 |
| **Mask Design** |  |  |  |  |  |  |  |  |  |  |  |
| **No-mask | 14 | 5 | 100.0% | --<sup>a</sup> | -- | 1.059<br>$\times 10^6$ | 2.237<br>$\times 10^5$ | 0.092 | 0.084 | --<sup>a</sup> | -- |
| Bandana style | 16 | 1 | 100.0% | 0.113 | 0.123 | 1.709<br>$\times 10^6$ | 9.205<br>$\times 10^5$ | 0.435 | 0.142 | 0.983 | 0.020 |
| Folded No-sew | 16 | 1 | 83.3% | 0.176 | 0.166 | 1.772<br>$\times 10^6$ | 7.754<br>$\times 10^5$ | 0.484 | 0.085 | 0.970 | 0.040 |
| Simple mask (short with earloops) | 16 | 1 | 100.0% | 0.119 | 0.159 | 1.651<br>$\times 10^6$ | 9.351<br>$\times 10^5$ | 0.263 | 0.175 | 0.981 | 0.035 |
| Stylistic mask | 16 | 1 | 100.0% | 0.208 | 0.240 | 1.839<br>$\times 10^6$ | 6.844<br>$\times 10^5$ | 0.341 | 0.127 | 0.968 | 0.050 |
| Surgical Mask style (internal non-woven) | 16 | 5 | 100.0% | 0.040 | 0.017 | 7.721<br>$\times 10^5$ | 6.606<br>$\times 10^5$ | 0.468 | 0.158 | 0.993 | 0.005 |
| **Exhalation Level** |  |  |  |  |  |  |  |  |  |  |  |
| Coughing | 48 | 1 | 100.0% | 0.154 | 0.158 | 1.388<br>$\times 10^6$ | 8.291<br>$\times 10^5$ | 0.353 | 0.191 | 0.988 | 0.013 |
| Talking | 46 | 5 | 66.7% | 0.104 | 0.151 | 1.567<br>$\times 10^6$ | 8.333<br>$\times 10^5$ | 0.352 | 0.186 | 0.971 | 0.042 |

a. The test condition for “no-mask” resulted in particle cloud velocities that exceeded the sampling rate of the PMS sensors and we not captured. Consequently, an EDF for “no-mask” was not calculated. From existing research, peak exhalation speeds can reach up to 10-30 m/s at the mouth[8] with expected slowdown at further distances.

**Figure 2:**
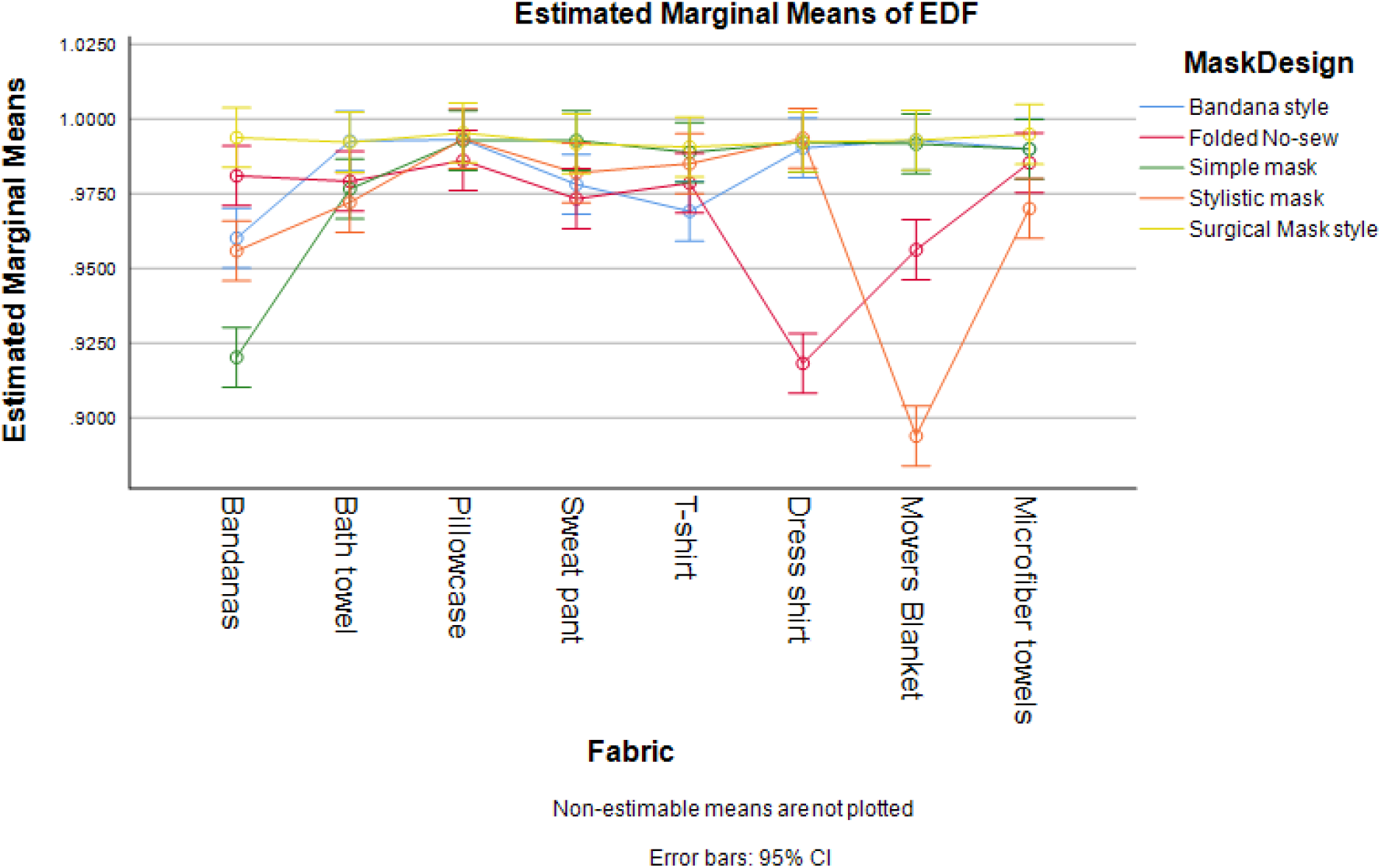
Estimated means of EDF resulting from MANOVA, showing the combined effects of fabrics and mask design. The test condition for “no-mask” resulted in particle cloud velocities that exceeded the sampling rate of the PMS sensors and were not captured. Consequently, an EDF for “no-mask” was not calculated.

The results from MANOVA on the complete data set are reported in Table 3 with all of the primary factors of fabric, mask design, and exhalation breath level resulting in an overall statistically significant effect on the dependent variables of direction, velocity, AUC_0 to 1 min_, FEI and EDF (Fabric: *F*(35, 36) = 8.526, *P* = <.001, Wilks' Λ = .000, Partial *η*^2^ = 846; Mask design: *F*(20, 27) = 15.691, *P* = <.001, Wilks' Λ = .000, Partial *η*^2^ =.876; Breath level: *F*(5, 8) = 371.6, *P* = <.001, Wilks' Λ = .004, Partial *η*^2^ =.996). Similarly, Table 3 shows that the results using Pillai’s Trace are also significant (in case the MANOVA assumptions of homogeneity of variance-covariance were violated). Therefore, the null hypothesis that masks or face coverings have no effect on exhalation dispersion or source control is rejected.

**Table 3.** Multivariate Tests showing significant factors affecting direction, velocity, AUC, FEI and EDF.^a^

| Effect | | Value | F | df | Error df | P value | Partial $\eta^2$ | Noncentrality Parameter |
| --- | --- | --- | --- | --- | --- | --- | --- | --- |
| **Intercept** | Pillai's Trace | 1.000 | 4.12x10<sup>6b</sup> | 5.000 | 8.000 | < .001 | 1.000 | 20.618x10<sup>6</sup> |
|  | Wilks' Lambda | 0.000 | 4.12x10<sup>6b</sup> | 5.000 | 8.000 | < .001 | 1.000 | 20.618x10<sup>6</sup> |
| **Fabric** | Pillai's Trace | 3.243 | 3.164 | 35.000 | 60.000 | < .001 | 0.649 | 110.731 |
|  | Wilks' Lambda | 0.000 | 8.526 | 35.000 | 36.083 | < .001 | 0.846 | 198.845 |
| **MaskDesign** | Pillai's Trace | 2.879 | 5.650 | 20.000 | 44.000 | < .001 | 0.720 | 113.009 |
|  | Wilks' Lambda | 0.000 | 15.691 | 20.000 | 27.483 | < .001 | 0.876 | 194.446 |
| **BreathLevel** | Pillai's Trace | 0.996 | 371.599<sup>b</sup> | 5.000 | 8.000 | < .001 | 0.996 | 1857.993 |
|  | Wilks' Lambda | 0.004 | 371.599<sup>b</sup> | 5.000 | 8.000 | < .001 | 0.996 | 1857.993 |
| **Fabric \* MaskDesign** | Pillai's Trace | 4.420 | 3.269 | 140.000 | 60.000 | < .001 | 0.884 | 457.631 |
|  | Wilks' Lambda | 0.000 | 6.925 | 140.000 | 44.549 | < .001 | 0.954 | 930.323 |
| **Fabric \* BreathLevel** | Pillai's Trace | 3.355 | 3.497 | 35.000 | 60.000 | < .001 | 0.671 | 122.397 |
|  | Wilks' Lambda | 0.000 | 7.820 | 35.000 | 36.083 | < .001 | 0.836 | 184.155 |
| **MaskDesign \* BreathLevel** | Pillai's Trace | 2.857 | 5.496 | 20.000 | 44.000 | < .001 | 0.714 | 109.923 |
|  | Wilks' Lambda | 0.001 | 10.703 | 20.000 | 27.483 | < .001 | 0.835 | 139.136 |
| **Fabric\* MaskDesign \* BreathLevel** | Pillai's Trace | 4.390 | 3.083 | 140.000 | 60.000 | < .001 | 0.878 | 431.560 |
|  | Wilks' Lambda | 0.000 | 5.729 | 140.000 | 44.549 | < .001 | 0.945 | 771.142 |

a. Design:
Intercept + Fabric + MaskDesign + BreathLevel + Fabric \* MaskDesign + Fabric \* BreathLevel + MaskDesign \* BreathLevel + Fabric \* MaskDesign \* BreathLevel
b. Exact statistic

The statistics of Wilks Lambda and Pillai’s trace (Table 3) converge at *η*^2^ = .996 and indicate that 99.6% of the variance of dependent variables are associated with exhalation breath levels of talking or coughing. It should also be noted that there were statistically significant interaction effects between fabric, mask design, breath levels and the combination of all three independent variables also reported in Table 3 (Fabric*Mask Design, Fabric*Breath level, Mask design*Breath level, Fabric*Mask design*Breath level). In some cases, the between-subject effects were marginally significant (P-value closer to .05), however the vast majority of individual between-subject effects 30/35 (85.7%) are significant. A full multivariate analysis of variance and multivariate tests of between-subject effects and interactions are provided in (online Supplementary Table 2 and 3).

## DISCUSSION

Conclusively, this quantitative effectiveness study establishes that improvised non-medical grade mask designs or fabric combinations are statistically significant in reducing airborne dispersion of particles from exhalation as defined by direction, velocity, AUC_0 to 1 min_, FEI and EDF. The statistically significant interaction effects between combinations of all primary factors and partial *η*^2^ values further establish the strong correlation of outcomes to fabrics used, mask design, and exhalation breath levels. This foundational research offers an orthogonal but complimentary result to previous research on respiratory protection and personal protective equipment which exclusively looks at inhalation filtration and airflow pressure gradients that support respiration.

When considering airborne dispersion control (also known as source control in some literature) it is important to understand the primary mechanisms that affect the dispersion. The field of filtration theory offers significant understanding with the primary mechanisms for the respiratory use case: Interception, Brownian Diffusion, Inertial Impaction, and Sieving or blocking filtration.[55-58] Since the fibers and fabric meshes are typically larger than small aerosols or infectious particles (5μm diameters or smaller), the first three filtration mechanisms are most applicable to this study and other studies in the field of personal protective equipment (PPE). Special emphasis is placed on inertial impaction and Brownian diffusion to disrupt the velocity and direction of airborne dispersion. Further discussion on filtration mechanisms is provided in (online Supplementary Appendix).

One observation from this study is that the effectiveness in dispersion control varies greatly between the specific fabrics and mask design combinations. For example, the factors of fabric, mask design, and the interaction of fabric and exhalation breath level that have significant effect on FEI, while other interactions are not significant (online Supplementary Table 3). This suggests that a fabric’s dynamic characteristics such as pliability (i.e. conforms to the face for fit and coverage) and dynamic response to airflow force (i.e. stretch characteristics) have an effect on the overall filtration of exhaled particles. Ad hoc test data of a stretch fabric commercially available mask is consistent with this statement (online Supplementary Table 5). Further materials analysis and characterization is justified to fully understand this observation however it does emphasize that proper wearing of masks fully covering the mouth and nose[59-61] is important for PPE usage as well as dispersion control. Characterization of fabric thickness, fiber density and weave, as well as layering will also aid in establishing accurate predictor coefficients of a dispersion control linear regression model for specific fabrics and masks.

The strength of this study is that it offers quantitative evidence on the effectiveness of non-medical improvised masks for helping to establishing public health strategies or policies that encourage the wearing of masks or face coverings. Fundamentally the effectiveness of non-pharmaceutical interventions (NPI) can be increased by reducing exhalation particle dispersion and is especially important where infectious contaminants may exist in shared air spaces. Universal application of masks will reduce overall particle quantities and slow down the dispersion speed which gives people in the vicinity of the dispersion (from coughing or talking) time to react and avoid inhalation exposure to potentially infectious particle clouds. The slowing down of a particle cloud’s dispersion also means that social distances will be more effective considering the relationship of velocity, time, and distance. This concept is well aligned with typical hazardous materials and bio-chem emergency response considerations of “time-distance-shielding” [62-64] where masks provide shielding during inhalation, and also slows exhalation particles to give others time and distance to avoid the particle cloud.

An overall public health strategy must consider the additive effect of wearing masks and face coverings for inhalation filtration (PPE) and that of dispersion and source control. However, the strategy would also need to account for the non-ideal performance of various fabrics and masks, where the ideal performance would offer 95% filtration efficiencies and dispersion contained to the user’s body. Combining this research with recent community SIR modeling[65] can help provide significant insights to the public health strategy. To summarize, it would be of most benefit for all people in community settings to wear masks and get full effect of controlling exhalation particle dispersion to reduce transmission of highly infectious respiratory diseases such as COVID-19.

### Limitations

One limitation of this study is that it provides approximations of human exhalation using polydisperse NaCl solution rather than human exhalation which adds additional compositions of particles that can be smaller than 0.3μm in diameters, moisture, proteins, gases, and other bio material.[66] The effectiveness of masks for source or dispersion control from long term use (such as during a 4 hour or 8 hour workdays) cannot be directly established from this data, however this study utilizes industry and NIOSH accepted proxy for testing respiratory barriers of NaCl. Another limitation was the PMS sensor performance: measurement minimum of 0.3μm particles and a slower intake fan speed at 0.1 CFM limited its ability to accurately measure all characteristics of fast moving particle clouds from that of no-mask applied. Regardless, the sensor data and experiment design were sufficient to determine statistical conclusions on the effects of wearing masks and face coverings of different fabrics and designs. Future works should consider using a large test chamber and more sensors to result in more accurate measurement of airborne dispersion and turbulent airflows.

## CONCLUSIONS

The results show that the application of various non-medical grade mask designs or fabric combinations were statistically significant in reducing airborne dispersion of particles from exhalation during coughing and talking as well as singing. However, the effectiveness varies greatly between the specific fabrics and mask designs used. The best overall performing mask design is a surgical style with internal non-woven layers, while the best overall fabric depends on a desired characteristic of reduced velocity, change in direction, or increased filtration performance. Conclusively this study can aid in establishing public health strategies or policies that encourage the wearing of masks or face coverings to increase the effectiveness of non-pharmaceutical interventions (NPI) especially where infectious contaminants may exist in shared air spaces.

## Data Availability

Empirical data may be made available by formal request to author's organization

## AUTHOR CONTRIBUTIONS

NE was the principal investigator and primary author, RW created the design of experiment and conducted the mathematical analysis, RP conducted background research and guided the approach to experimentation and rigor of analysis, MG assisted in the laboratory configuration and testing.

## ACKNOWLEDGMENT

We extend our appreciation to The American Red Cross of Southeastern Colorado and El Paso County Public Health for allowing the use of CPR training equipment and manikin to rapidly conduct this study. We also acknowledge Mr. Asher Edwards for his contributions to the early code development for the data acquisition system, and Ms. Lydia Edwards for providing continuous awareness of open news research and public vetting of concepts.

## DATA AVAILABILITY STATEMENT

The datasets generated from this study are available to U.S. national, state and regional agencies as well as healthcare systems on request to the corresponding author.

## REFERENCES

1. van Doremalen N, Bushmaker T, Morris DH, Holbrook MG, Gamble A, Williamson BN, et al. Aerosol and Surface Stability of SARS-CoV-2 as Compared with SARS-CoV-1. N Engl J Med. 2020;382:1564–7.

2. Milton DK, Fabian MP, Cowling BJ, Grantham ML, McDevitt JJ. Influenza Virus Aerosols in Human Exhaled Breath: Particle Size, Culturability, and Effect of Surgical Masks. PLOS Pathog. 2013;9:e1003205.

3. Leung NHL, Chu DKW, Shiu EYC, Chan K-H, McDevitt JJ, Hau BJP, et al. Respiratory virus shedding in exhaled breath and efficacy of face masks. Nat Med. 2020;26:676–80.

4. Asadi S, Wexler AS, Cappa CD, Barreda S, Bouvier NM, Ristenpart WD. Aerosol emission and superemission during human speech increase with voice loudness. Sci Rep. 2019;9:1–10.

5. Stadnytskyi V, Bax CE, Bax A, Anfinrud P. The airborne lifetime of small speech droplets and their potential importance in SARS-CoV-2 transmission. Proc Natl Acad Sci. 2020;117:11875–7.

6. Papineni RS, Rosenthal FS. The Size Distribution of Droplets in the Exhaled Breath of Healthy Human Subjects. J Aerosol Med. 1997;10:105–16.

7. Morawska L, Johnson GR, Ristovski ZD, Hargreaves M, Mengersen K, Corbett S, et al. Size distribution and sites of origin of droplets expelled from the human respiratory tract during expiratory activities. J Aerosol Sci. 2009;40:256–69.

8. Bourouiba L. Turbulent Gas Clouds and Respiratory Pathogen Emissions: Potential Implications for Reducing Transmission of COVID-19. JAMA. 2020. doi:10.1001/jama.2020.4756.

9. Pan M, Lednicky JA, Wu C-Y. Collection, particle sizing and detection of airborne viruses. J Appl Microbiol. 2019;127:1596–611.

10. Chu DK, Akl EA, Duda S, Solo K, Yaacoub S, Schünemann HJ, et al. Physical distancing, face masks, and eye protection to prevent person-to-person transmission of SARS-CoV-2 and COVID-19: a systematic review and meta-analysis. The Lancet. 2020;395:1973–87.

11. Atkinson J, World Health Organization, editors. Natural ventilation for infection control in health-care settings. Geneva: World Health Organization; 2009. https://www.ncbi.nlm.nih.gov/books/NBK143284/.

12. Tellier R, Li Y, Cowling BJ, Tang JW. Recognition of aerosol transmission of infectious agents: a commentary. BMC Infect Dis. 2019;19:101.

13. MacIntyre CR, Cauchemez S, Dwyer DE, Seale H, Cheung P, Browne G, et al. Face Mask Use and Control of Respiratory Virus Transmission in Households. Emerg Infect Dis. 2009;15:233–41.

14. Mukerji S, MacIntyre CR, Newall AT. Review of economic evaluations of mask and respirator use for protection against respiratory infection transmission. BMC Infect Dis. 2015;15:413.

15. National Academies of Sciences E. Rapid Expert Consultation on the Effectiveness of Fabric Masks for the COVID-19 Pandemic (April 8, 2020). 2020. doi:10.17226/25776.

16. Aiello AE, Murray GF, Perez V, Coulborn RM, Davis BM, Uddin M, et al. Mask use, hand hygiene, and seasonal influenza-like illness among young adults: A randomized intervention trial. J Infect Dis. 2010;201:491–8.

17. Agah R, Cherry JD, Garakian AJ, Chapin M. Respiratory Syncytial Virus (RSV) Infection Rate in Personnel Caring for Children With RSV Infections: Routine Isolation Procedure vs Routine Procedure Supplemented by Use of Masks and Goggles. Am J Dis Child. 1987;141:695–7.

18. Relman E. Trump shares tweet that says masks represent “slavery and social death” - Business Insider. 2020. https://www.businessinsider.com/trump-shares-tweet-that-says-masks-represent-slavery-and-social-death-2020-5. Accessed 28 May 2020.

19. McCann M. Mandatory Masks Aren’t About Safety, They’re About Social Control. 2020. https://thefederalist.com/2020/05/27/mandatory-masks-arent-about-safety-theyre-about-social-control/. Accessed 28 May 2020.

20. Chuck E, Einhorn E, Gamboa S, Hassan A, Kingkade T, Smith S. The mask wearing debate is dividing America. And the messaging isn’t getting any clearer. MSN. 2020. https://www.msn.com/en-us/news/us/the-mask-wearing-debate-is-dividing-america-and-the-messaging-isnt-getting-any-clearer/ar-BB15BxDd. Accessed 19 Jun 2020.

21. Brooks JT, Butler JC, Redfield RR. Universal Masking to Prevent SARS-CoV-2 Transmission—The Time Is Now. JAMA. 2020. doi:10.1001/jama.2020.13107.

22. Wang X, Ferro EG, Zhou G, Hashimoto D, Bhatt DL. Association Between Universal Masking in a Health Care System and SARS-CoV-2 Positivity Among Health Care Workers. JAMA. 2020. doi:10.1001/jama.2020.12897.

23. Fisher KA. Factors Associated with Cloth Face Covering Use Among Adults During the COVID-19 Pandemic — United States, April and May 2020. MMWR Morb Mortal Wkly Rep. 2020;69. doi:10.15585/mmwr.mm6928e3.

24. Melinda Mills, Charles Rahal, Evelina Akimova. Face masks and coverings for the general public: Behavioural knowledge, effectiveness of cloth coverings and public messaging. The Royal Society; 2020. https://royalsociety.org/topics-policy/projects/set-c-science-in-emergencies-tasking-covid/.

25. Hendrix MJ, Charles Walde, Kendra Findley, Robin Trotman. Absence of Apparent Transmission of SARS-CoV-2 from Two Stylists After Exposure at a Hair Salon with a Universal Face Covering Policy — Springfield, Missouri, May 2020. MMWR Morb Mortal Wkly Rep. 2020;69. doi:10.15585/mmwr.mm6928e2.

26. Mueller AV, Fernandez LA. Assessment of Fabric Masks as Alternatives to Standard Surgical Masks in Terms of Particle Filtration Efficiency. medRxiv. 2020;:2020.04.17.20069567.

27. Nallathambi G, S E, R K, D N. Multilayer nonwoven fabrics for filtration of micron and submicron particles. J Text Eng Fash Technol. 2019;5. doi:10.15406/jteft.2019.05.00185.

28. Rengasamy S, Eimer B, Shaffer RE. Simple respiratory protection--evaluation of the filtration performance of cloth masks and common fabric materials against 20-1000 nm size particles. Ann Occup Hyg. 2010;54:789–98.

29. Konda A, Prakash A, Moss GA, Schmoldt M, Grant GD, Guha S. Aerosol Filtration Efficiency of Common Fabrics Used in Respiratory Cloth Masks. ACS Nano. 2020;14:6339–47.

30. Shakya KM, Noyes A, Kallin R, Peltier RE. Evaluating the efficacy of cloth facemasks in reducing particulate matter exposure. J Expo Sci Environ Epidemiol. 2017;27:352–7.

31. Michigan Mask Response. DIY Mask FAQ - Test Results. DIY Mask FAQ. https://www.maskfaq.com/test-results. Accessed 6 May 2020.

32. Robertson P. The Ultimate Guide to Homemade Face Masks for Coronavirus. Smart Air Filters. 2020. https://smartairfilters.com/en/blog/best-diy-coronavirus-homemade-mask-material-covid/. Accessed 22 Apr 2020.

33. Lustig SR, Biswakarma JJH, Rana D, Tilford SH, Hu W, Su M, et al. Effectiveness of Common Fabrics to Block Aqueous Aerosols of Virus-like Nanoparticles. ACS Nano. 2020;14:7651–8.

34. Verma S, Dhanak M, Frankenfield J. Visualizing the effectiveness of face masks in obstructing respiratory jets. Phys Fluids. 2020;32:061708.

35. Lu J, Gu J, Li K, Xu C, Su W, Lai Z, et al. COVID-19 Outbreak Associated with Air Conditioning in Restaurant, Guangzhou, China, 2020. Emerg Infect Dis J. 2020;26:1628.

36. Dietz L, Horve PF, Coil DA, Fretz M, Eisen JA, Van Den Wymelenberg K. 2019 Novel Coronavirus (COVID-19) Pandemic: Built Environment Considerations To Reduce Transmission. mSystems. 2020;5:e00245-20, /msystems/5/2/msys.00245-20.atom.

37. Blocken B, Malizia F, van Druenen T, Marchal T. Towards aerodynamically equivalent COVID19 1.5 m social distancing for walking and running.:12.

38. Grush L. How Sneeze Particles Travel Inside An Airplane | Popular Science. 2014. https://www.popsci.com/article/science/how-sneeze-particles-travel-inside-airplane/. Accessed 15 Jun 2020.

39. Gröndahl M, Goldbaum C, White J. What Happens to Viral Particles on the Subway. The New York Times. 2020. https://www.nytimes.com/interactive/2020/08/10/nyregion/nyc-subway-coronavirus.html. Accessed 10 Aug 2020.

40. Davies A, Thompson K-A, Giri K, Kafatos G, Walker J, Bennett A. Testing the Efficacy of Homemade Masks: Would They Protect in an Influenza Pandemic? Disaster Med Public Health Prep. 2013;7:413–8.

41. Patel RB, Skaria SD, Mansour MM, Smaldone GC. Respiratory source control using a surgical mask: An in vitro study. J Occup Environ Hyg. 2016;13:569–76.

42. Abd-Elsayed A, Karri J. Utility of Substandard Face Mask Options for Health Care Workers During the COVID-19 Pandemic. Anesth Analg. 2020;131:4–6.

43. Eikenberry SE, Mancuso M, Iboi E, Phan T, Eikenberry K, Kuang Y, et al. To mask or not to mask: Modeling the potential for face mask use by the general public to curtail the COVID-19 pandemic. Infect Dis Model. 2020;5:293–308.

44. Lindsley WG, King WP, Thewlis RE, Reynolds JS, Panday K, Cao G, et al. Dispersion and Exposure to a Cough-Generated Aerosol in a Simulated Medical Examination Room. J Occup Environ Hyg. 2012;9:681–90.

45. Lindsley WG, Reynolds JS, Szalajda JV, Noti JD, Beezhold DH. A Cough Aerosol Simulator for the Study of Disease Transmission by Human Cough-Generated Aerosols. Aerosol Sci Technol. 2013;47:937–44.

46. Hui DS, Chow BK, Chu L, Ng SS, Lee N, Gin T, et al. Exhaled air dispersion during coughing with and without wearing a surgical or N95 mask. PloS One. 2012;7:e50845.

47. Rothenberg M, Miller D, Molitor R, Leffingwell D. The control of air flow during loud soprano singing. J Voice. 1987;1:262–8.

48. National Institute for Occupational Safety and Health. TEB-APR-STP-0059: Determination of Particulate Filter Efficiency Level for N95 Series Filters Against Solid Particulates for Non-powered, Air-purifying Respirators Standard Testing Procedure (STP). 2019. https://www.cdc.gov/niosh/npptl/stps/pdfs/TEB-APR-STP-0059-508.pdf.

49. Ower E, Pankhurst RC. The Measurement of Air Flow. Elsevier; 2014.

50. Warne R. A Primer on Multivariate Analysis of Variance (MANOVA) for Behavioral Scientists. 2014;19:11.

51. Ito PK. 7 Robustness of ANOVA and MANOVA test procedures. In: Handbook of Statistics. Elsevier; 1980. p. 199-236. doi:10.1016/S0169-7161(80)01009-7.

52. Handbook of Applied Multivariate Statistics and Mathematical Modeling. Elsevier; 2000. doi:10.1016/B978-0-12-691360-6.X5000-9.

53. Huberty CJ, Petoskey MD. 7 - Multivariate Analysis of Variance and Covariance. In: Tinsley HEA, Brown SD, editors. Handbook of Applied Multivariate Statistics and Mathematical Modeling. San Diego: Academic Press; 2000. p. 183-208. doi:10.1016/B978-012691360-6/50008-2.

54. Crichton N. Information Point: Wilks’ lambda. J Clin Nurs. 2000;:369-81.

55. Schmidt EW, Gieseke JA, Gelfand P, Lugar TW, Furlong DA. Filtration Theory for Granular Beds. J Air Pollut Control Assoc. 1978;28:143–6.

56. Ranz WE, Katz EJ. Determining Impaction Efficiencies of Mist Collection Equipment. J Air Pollut Control Assoc. 1959;8:328–32.

57. Ripperger S, Gösele W, Alt C. Filtration, 1. Fundamentals. In: Wiley-VCH Verlag GmbH & Co. KGaA, editor. Ullmann’s Encyclopedia of Industrial Chemistry. Weinheim, Germany: Wiley-VCH Verlag GmbH & Co. KGaA; 2009. p. b02_10.pub2. doi:10.1002/14356007.b02_10.pub2.

58. Sipes J. BASICS OF AIR FILTRATION. Price Industries. https://www.priceindustries.com/content/uploads/assets/literature/technical-papers/basics-of-air-filtration.pdf.

59. Desai AN, Aronoff DM. Masks and Coronavirus Disease 2019 (COVID-19). JAMA. 2020;323:2103–2103.

60. CDC. Recommendation Regarding the Use of Cloth Face Coverings, Especially in Areas of Significant Community-Based Transmission. Centers for Disease Control and Prevention. 2020. https://www.cdc.gov/coronavirus/2019-ncov/prevent-getting-sick/cloth-face-cover.html. Accessed 3 Apr 2020.

61. CDC. Use of Cloth Face Coverings to Help Slow the Spread of COVID-19. Centers for Disease Control and Prevention. 2020. https://www.cdc.gov/coronavirus/2019-ncov/prevent-getting-sick/diy-cloth-face-coverings.html. Accessed 16 Apr 2020.

62. Robert Avsec. Time, distance, shielding: Minimizing radiation exposure. FireRescue1. 2019. https://www.firerescue1.com/fire-products/hazmat-equipment/articles/time-distance-shielding-minimizing-radiation-exposure-yUjpcxrDyYFdibTa/. Accessed 12 Aug 2020.

63. U.S. Department of Health and Human Services. Chemical Hazards Emergency Medical Management - Quick Reponse Guide. 2020. https://chemm.nlm.nih.gov/quickresponseguide.htm. Accessed 12 Aug 2020.

64. U.S. Department of Health and Human Services. The Golden First Minutes — Initial Response to a Chemical Hazardous Materials Incident - CHEMM. 2020. https://chemm.nlm.nih.gov/detailedinfo.htm. Accessed 12 Aug 2020.

65. Stutt ROJH, Retkute R, Bradley M, Gilligan CA, Colvin J. A modelling framework to assess the likely effectiveness of facemasks in combination with ‘lock-down’ in managing the COVID-19 pandemic. Proc R Soc Math Phys Eng Sci. 2020;476:20200376.

66. Popov TA. Human exhaled breath analysis. Ann Allergy Asthma Immunol. 2011;106:451–6.

